# Immune Markers Mediate Genetic Relationships between Immune Diseases and Psychiatric Disorders

**DOI:** 10.1101/2025.10.17.25338241

**Authors:** Sophie Breunig, Kristen M. Kelly, Lukas S. Schaffer, Jeremy M. Lawrence, Younga Heather Lee, Elizabeth W. Karlson, Andrew D. Grotzinger

## Abstract

This study explored the genetic links between 11 immune-mediated diseases and 13 psychiatric disorders, addressing the limited understanding of what links these illnesses. Utilizing genomic structural equation modeling, we investigated whether 14 immune markers (e.g., C-reactive protein [CRP], interleukin-6 [IL-6]) mediated associations between latent genomic factors of immune-mediated diseases (e.g., autoimmune) and psychiatric disorders (e.g., internalizing). While the literature extensively describes the psychiatric relevance of CRP, results indicated this is better explained by its biological precursor, IL-6. Six immune markers significantly mediated 11 psychiatric-immune disease genetic relationships. For example, IL-6 mediated 60% of the genetic link between an internalizing factor and Crohn’s disease. Identified markers often evinced mediating effects for multiple disorder clusters, while at the same time showing some specificity in their associations. These findings provide mechanistic insight by pinpointing the immune markers that may be most critical to the psychiatric-immune disease link, which notably did not include CRP.

---

Autoimmune and inflammatory diseases, collectively referred to as immune-mediated diseases, are conditions in which an abnormal immune response leads to prolonged activation of immune and inflammatory pathways. A growing body of literature describes high levels of comorbidity across a wide range of immune-mediated diseases and psychiatric disorders^1–3^. In line with the pervasive diagnostic overlap observed at the phenotypic level, recent genomic findings revealed that genetic overlap across an individual psychiatric disorder and immune-mediated diseases often indexed broader pathways of risk sharing^4^. For example, while schizophrenia and bipolar disorder have each been separately linked to ulcerative colitis and Crohn’s disease^5–7^, these risk pathways have now been shown to be shared across all four disorders^4^. The current study aims to build on this prior work to broaden our understanding of possible intermediate links between immune-mediated diseases and psychiatric disorders.

Immune and inflammatory markers are plausible mediating mechanisms given prior findings linking these markers to both immune diseases and psychiatric disorders. With respect to immune-mediated disease, these disease states are characterized by chronic activation of the immune system that is reflected in the relative increase or decrease of certain markers of immune and inflammatory activity^8–10^. An extensive prior literature has described these links between these biomarkers and several immune-mediated diseases. For example, tumor necrosis factor alpha (TNF-α) has been found to be positively genetically correlated with a host of different immune-mediated diseases^11^. These markers have also been widely associated with psychiatric disorders. This includes relationships described between interleukin-1 (IL-1), interleukin-6 (IL-6) and C-reactive protein (CRP) and increased levels in depression in a dose-response fashion, where greater levels of the markers were associated with greater severity of depression symptoms^12^. Altered levels of immune markers have been reported for a wide range of psychiatric disorders, including generalized anxiety disorder^13^, schizophrenia^14^, bipolar disorder^15^, and attention-deficit/hyperactivity disorder (ADHD)^16^.

Convergent lines of evidence linking psychiatric disorders, immune-mediated diseases, and immune markers indicate that critical knowledge can be gained by jointly examining these three sets of outcomes. A limiting factor for this specific line of questioning reflects pragmatic difficulties in collecting a participant sample that has enough observations for even a single psychiatric disorder, immune-mediated disease, and immune marker. Here, we utilize the unique abilities of genetic data to bring together non-overlapping datasets by examining genetic overlap at the level of the effects of genetic variants. We specifically apply a multivariate genomic approach, genomic structural equation modeling (genomic SEM), and its extensions to evaluate whether a comprehensive set of immune markers mediates genetic relationships between immune and psychiatric disorders.

## Results

### Psychiatric and Immune Factor Structure

The current analyses utilized genome-wide association study (GWAS) summary statistics of 11 immune-mediated diseases, 13 psychiatric disorders, and 14 immune markers (**Table 1**). We began by using Genomic SEM to model the factor structure, based on genetic correlations derived from linkage disequilibrium score regression (LDSC), of the psychiatric disorders and immune diseases.

**Table 1.** Sample Characteristics of Included Traits.

| Phenotype | Cases | Controls | Liability Scale Heritability |
| --- | --- | --- | --- |
| *Immune Markers* |  |  |  |
| C-C motif chemokine 2 (CCL2)<sup>37</sup> | 46 644 | - | 8% |
| C-C motif chemokine 3 (CCL3)<sup>37</sup> | 46 643 | - | 15% |
| C-reactive protein (CRP)<sup>38</sup> | 427 367 | - | 15% |
| Interferon gamma (IFN-γ)<sup>37</sup> | 46 643 | - | 5% |
| Interleukin 10 (IL-10)<sup>37</sup> | 46 643 | - | 5% |
| Interleukin 1 beta (IL-1β)<sup>37</sup> | 46 643 | - | 5% |
| Interleukin 1 receptor antagonist (IL-1RA)<sup>37</sup> | 46 643 | - | 15% |
| Interleukin 6 (IL-6)<sup>37</sup> | 46 644 | - | 11% |
| Interleukin 6 receptor subunit alpha (IL-6RA)<sup>37</sup> | 46 643 | - | 8% |
| Soluble glycoprotein 130 (SGP130)<sup>37</sup> | 46 643 | - | 8% |
| Transforming growth factor beta-1 proprotein (TGF- β1)<sup>37</sup> | 46 643 | - | 12% |
| Tumor necrosis factor alpha (TNF-α)<sup>37</sup> | 46 644 | - | 9% |
| Tumor necrosis factor receptor superfamily member 1A (TNFR1A)<sup>37</sup> | 46 644 | - | 17% |
| Tumor necrosis factor receptor superfamily member 1B (TNFR1B)<sup>37</sup> | 46 644 | - | 18% |
| *Immune Mediated Diseases* |  |  |  |
| Addison's disease (AD)<sup>39</sup> | 1 223 | 4 097 | 16% |
| Celiac disease (CEL)<sup>40</sup> | 2 364 | 324 074 | 13% |
| Crohn's disease (CD)<sup>41</sup> | 12 194 | 28 072 | 22% |
| Diabetes type 1 (T1D)<sup>42</sup> | 18 942 | 501 638 | 14% |
| Juvenile idiopathic arthritis (JIA)<sup>43</sup> | 3 305 | 9 196 | 13% |
| Myasthenia gravis (MG)<sup>44</sup> | 1 873 | 36 370 | 11% |
| Psoriasis (PSO)<sup>45</sup> | 15 967 | 28 194 | 15% |
| Psoriatic arthritis (PSA)<sup>46</sup> | 5 065 | 21 286 | 13% |
| Rheumatoid arthritis (RA)<sup>47</sup> | 14 361 | 43 923 | 9% |
| Systemic lupus erythematosus (SLE)<sup>48</sup> | 4 036 | 6 959 | 10% |
| Ulcerative colitis (UC)<sup>41</sup> | 12 366 | 33 609 | 15% |
| *Psychiatric Disorders* |  |  |  |
| Alcohol use disorder (ALCH)<sup>49</sup> | 8 485 | 20 272 | 14% |
| Anorexia nervosa (AN)<sup>50</sup> | 16 992 | 55 525 | 16% |
| Anxiety disorder (ANX)<sup>51</sup> | 31 977 | 82 114 | 23% |
| Attention-deficit/hyperactivity disorder (ADHD)<sup>52</sup> | 38 691 | 186 843 | 18% |
| Autism spectrum disorder (ASD)<sup>53</sup> | 18 381 | 27 969 | 12% |
| Bipolar disorder (BIP)<sup>54</sup> | 56 287 | 781 022 | 12% |
| Cannabis use disorder (CUD)<sup>55</sup> | 14 808 | 343 726 | 19% |
| Major depressive disorder (MDD)<sup>56</sup> | 412 305 | 1 588 397 | 7% |
| Obsessive compulsive disorder (OCD)<sup>57</sup> | 2 688 | 7 037 | 30% |
| Opioid use disorder (OUD)<sup>58</sup> | 3 211 | 11 589 | 19% |
| Post traumatic stress disorder (PTSD)<sup>59</sup> | 141 479 | 1 113 329 | 5% |
| Schizophrenia (SCZ)<sup>60</sup> | 53 386 | 77 258 | 22% |
| Tourette's syndrome (TS)<sup>61</sup> | 4 819 | 9 488 | 22% |

*Note.* This table describes all GWAS summary statistics of all traits included in the analyses. Traits where only a sample size for cases and not controls is given, indicate that this trait had a continuous N.

The psychiatric factor structure reflected a previously established five-factor model^17^, but with updated GWAS data for bipolar disorder, major depression, and post-traumatic stress disorder (PTSD) (**Method**). We found that the five-factor model continued to fit the data well with the updated data (CFI = .97; SRMR = .10). The psychiatric factors can be approximately described as reflecting compulsive, schizophrenia/bipolar, neurodevelopmental, internalizing, and substance use disorders.

The immune-mediated disease factor structure consisted of a previously established four-factor model^4^ (CFI = .96; SRMR = .08), with the four factors reflecting autoimmune, celiac, mixed immune pattern (denoting a shared involvement of the adaptive and innate immune system), and autoinflammatory diseases (**Figure 1**).

**Figure 1.**
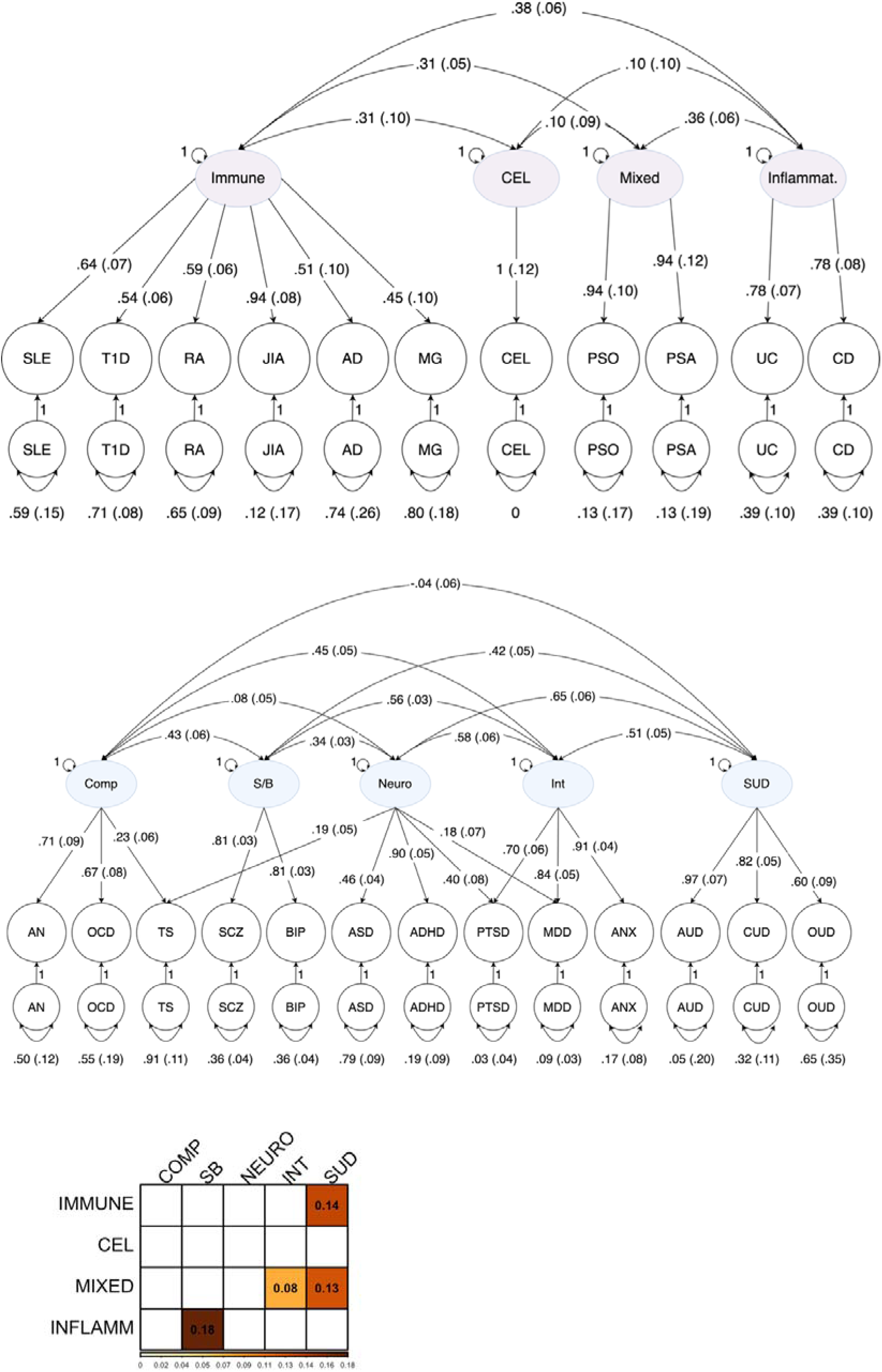
Immune-mediated Disease and Psychiatric Disorders Correlated Factors Models and Correlations between Immune and Psychiatric Factors. A) Immune-mediated diseases path diagram with standardized estimates for the correlated factors model used in genomic structural equation modelling (genomic SEM) with four factors of immune-mediated diseases. Latent variables are represented as circles. The genetic component of each phenotype is represented with a circle because the genetic component is a latent variable that is not directly measured but is inferred using linkage disequilibrium score regression (LDSC). Single-headed arrows are factor loadings; double- headed arrows connecting back to the same origin are variances; and double-headed arrows connecting two variables are correlations. Paths labeled 1 are fixed to 1; all estimates are given next to the path with standard errors in parentheses. Disease abbreviations are defined in Table 1. B) Psychiatric disorders path diagram. Disorder abbreviations are defined in Table 1. C) Genetic correlations between factors of immune-mediated diseases and factors of psychiatric disorders. IMMUNE, autoimmune diseases; CEL, celiac disease; MIXED, mixed immune pattern diseases; INFLAMM, inflammatory diseases; COMP, compulsive disorders; SB, schizophrenia/bipolar; NEURO, neurodevelopmental disorders; INT, internalizing disorders; SUD, substance use disorders.

The correlations across the psychiatric and immune factors were then estimated. Significance was defined using an FDR corrected threshold. We only considered inter-factor correlations significant when they were also not FDR significant for the *Q_Factor_* heterogeneity statistic, which was used as a quality control test to evaluate whether these factor correlations accurately captured genetic overlap^4^. Non-significant *Q_Factor_* results suggest that the factor correlation appropriately captures risk sharing across these two clusters of disease traits. Conversely, *Q_Factor_* would be expected to be significant if disorders loading on Factor 1 had a mixture of positive and negative genetic correlations with the disorders on Factor 2, such that a single correlation between Factor 1 and 2 is insufficient for describing the cross-disorder patterning of associations. Pruning out *Q_Factor_* significant associations yielded four significant inter-factor associations (**Supplemental Table S1**), reflecting correlations between autoimmune disease and substance use disorder factors, mixed immune pattern diseases with internalizing and substance use disorders factors, and inflammatory disorders with the schizophrenia/bipolar factor.

### Associations with Immune Markers

We next examined the genetic correlations between the 14 immune markers and the five psychiatric and four immune factors. We again used the *Q_Factor_* heterogeneity statistic to evaluate whether genetic correlations with the markers were likely to operate at the factor level. In instances where there were significant *Q_Factor_* results, we evaluated whether the immune marker was significantly associated with any of the individual disorders that defined the psychiatric or immune factor. After pruning out significant *Q_Factor_* results, we identified 20 FDR significant immune marker – immune disease relationships, which included associations of ten markers with three immune-disease factors (all but the celiac factor; **Supplemental Table S2**). Interferon gamma (IFNG) was the strongest associated marker for the inflammatory (*r_g_* = .48, standard error [*SE*] = .10) and mixed immune pattern factors (*r_g_* = .35, *SE* = .09), while TNF-α had the largest correlation for the immune factor (*r_g_* = .51, *SE* = .08). For individual disease associations investigated due to significant *Q_Factor_* results, we observed significant correlations between Crohn’s disease and CRP (*r_g_* = .16, *SE* = .03) and IL-6 (*r_g_* = .33, *SE* = .06), along with ulcerative colitis and IL-6 (*r_g_* = .12, *SE* = .06).

For the psychiatric factors, we identified 27 significant immune marker genetic correlations that included ten markers and four of the five psychiatric disorders factors (all but the schizophrenia/bipolar factor; **Supplemental Table S3**), along with five significant correlations with ADHD. Correlations were all positive, with the exception of large negative genetic correlations (range: -.21 : -.41) for 9 markers and the compulsive factor. Interleukin 1 receptor antagonist (IL- 1RA) had the largest point estimate across markers for the compulsive (*r_g_* = -.41, *SE* = .07) and internalizing factors (*r_g_* = .26, *SE* = .04) and for ADHD (*r_g_* = .35, *SE* = .04). C-C motif chemokine 3 (CCL3) had the largest correlation for the neurodevelopmental (*r_g_* = .23, *SE* = .05) and substance use factors (*r_g_* = .32, *SE* = .07).

Two pairs of markers, TNF receptor superfamily member 1A and B (TNFR1A & TNFR1B) and TNF and C-C motif chemokine 3 (CCL3), displayed highly similar patterns of genetic correlations across psychiatric and immune factors. Both marker pairs additionally consisted of highly genetically correlated (see **Supplemental Figure S1**) and mechanistically related markers in terms of their function within the immune system. TNFR1A and B are both representatives of the same TNF- receptor superfamily and part of the same signaling network that influences the effect of TNF-α on macrophage viability and is closely integrated with pattern recognition receptor regulated signaling systems^18^. Similarly, TNF-α and CCL3 are part of the same signaling pathway, with TNF-α directly promoting CCL3 production^19^. Consequently, these two marker pairs were each modeled as two-indicator factors in subsequent mediation analyses to reflect the joint genetic architecture, and previously described biology, that they share.

### Multiple Regression Models Reveal Null Effects of CRP

In line with prior work describing links between CRP and psychiatric conditions, significant correlations were found for the compulsive (*r_g_* = -.37, *SE* = .05), internalizing (*r_g_* = .13, *SE* = .02) and substance use disorder (*r_g_* = .23, *SE* = .04) factors and for ADHD (*r_g_* = .30, *SE* = .02). As IL-6 stimulates CRP production^20^, it is unsurprising that IL-6 showed a highly similar pattern of associations, again with the compulsive (*r_g_* = -.35, *SE* = .08), internalizing (*r_g_* = .23, *SE* = .04) and substance use disorder (*r_g_* = .29, *SE* = .07) factors and ADHD (*r_g_* = .34, *SE* = .05). Since CRP is produced by the liver and not able to cross the blood-brain-barrier (despite altering its permeability)^21^, and it is biologically more likely that IL-6 that can cross the blood-brain barrier^22^, it is plausible that CRP is merely correlated with psychiatric disorders due to its relationship with IL-6. To investigate this, we fit multiple regression models with CRP and IL-6 as correlated predictors to test the partial effects of each marker on the psychiatric disorders or immune diseases (**Supplemental Figure S2**). These revealed that only IL-6 continued to have significant partial associations (**Supplemental Table 4**). CRP was therefore dropped from all subsequent mediation models in favor of IL-6.

### Immune Markers Mediate Immune-Psychiatric Associations

We aimed to produce a robust set of findings by only testing mediation models that met three requirements: (i) the marker was significantly associated with an immune disease factor or disorder (often denoted the *a* – path in a mediation model), (ii) the marker was significantly associated with a psychiatric factor or disorder (*b* – path), and (iii) the disorders or factors from the *a* and *b* paths were themselves significantly associated (the total effect to be mediated, often denoted the *c* – path). This process resulted in three types of mediation models that tested a marker’s mediation of: a *factor-factor* relationship, representing an immune-mediated disease factor to psychiatric factor relationship; a *factor-trait* relationship, representing an immune-mediated disease factor to an individual psychiatric disorder relationship or a psychiatric disorder factor to and individual immune-mediated disease relationship, or finally; a *trait-trait* relationship that represents an individual immune-mediated disease to an individual psychiatric disorder relationship.

We tested 12 mediation models for six markers that met the inclusion criteria described above (as detailed in **Figure 2** with model fit statistics in **Supplemental Table 5** and model outputs in **Supplemental Tables 6-17**). Of these 12, 11 were significant at an FDR threshold. A graphical representation of the direct, indirect and total effects of each mediation is depicted in **Figure 3** and a pie chart of the proportions of these effects for each mediation model can be found in **Figure 4**. Six of the mediations were for *factor-factor* relationships, five were for *factor-trait* relationships. There were no *trait-trait* relationships that exhibited significant mediation by an immune marker.

**Figure 2.**
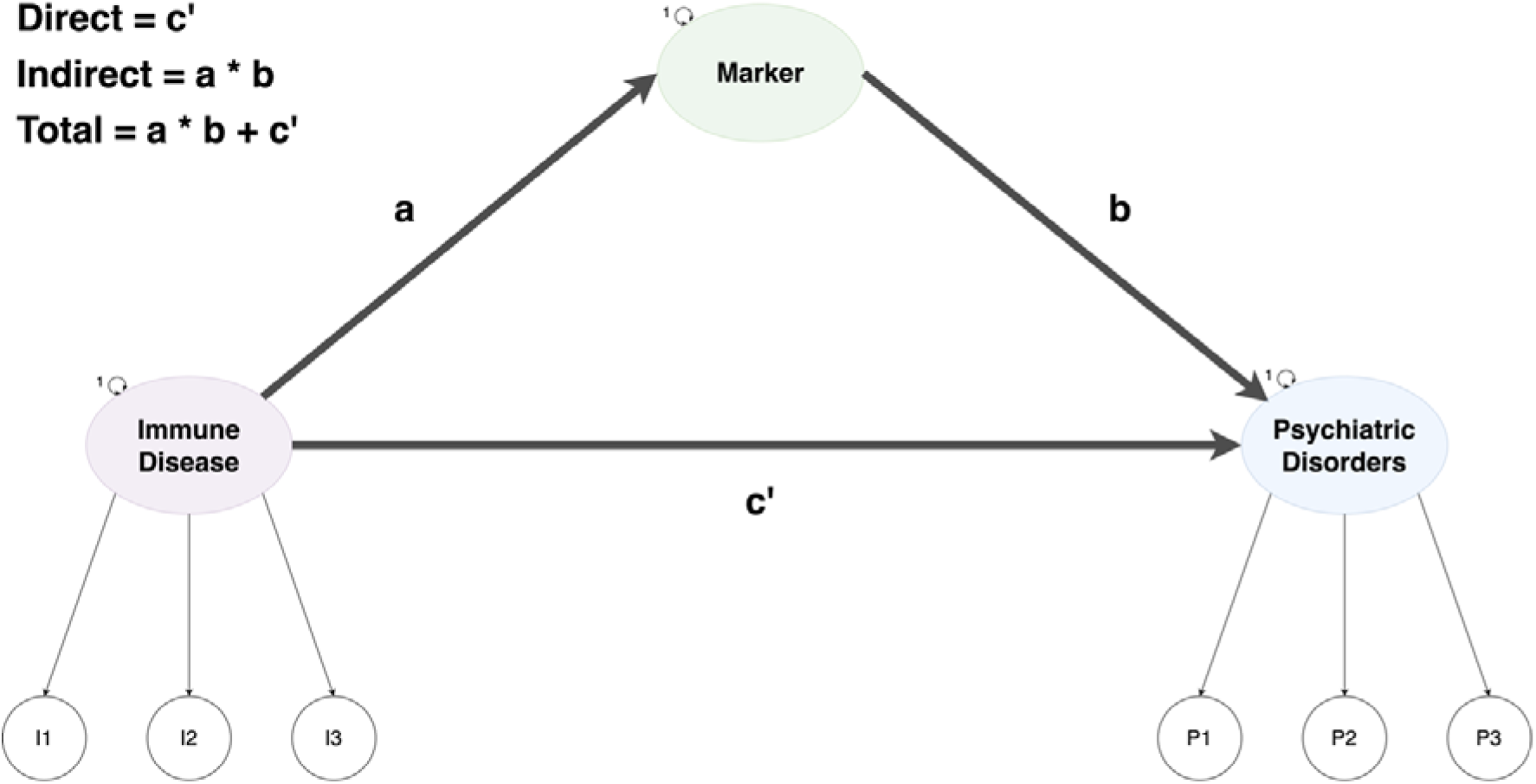
Example Mediation Model. Example of a mediation model for the relationship between an immune disease factor with indicators I1-3 and a psychiatric disorder factor with the indicators P1-3. The marker serves as a mediator and the corresponding *a*, *b*, and *c*’-path are tested and with them, the direct, indirect, and total effects are calculated according to the above equations.

**Figure 3.**
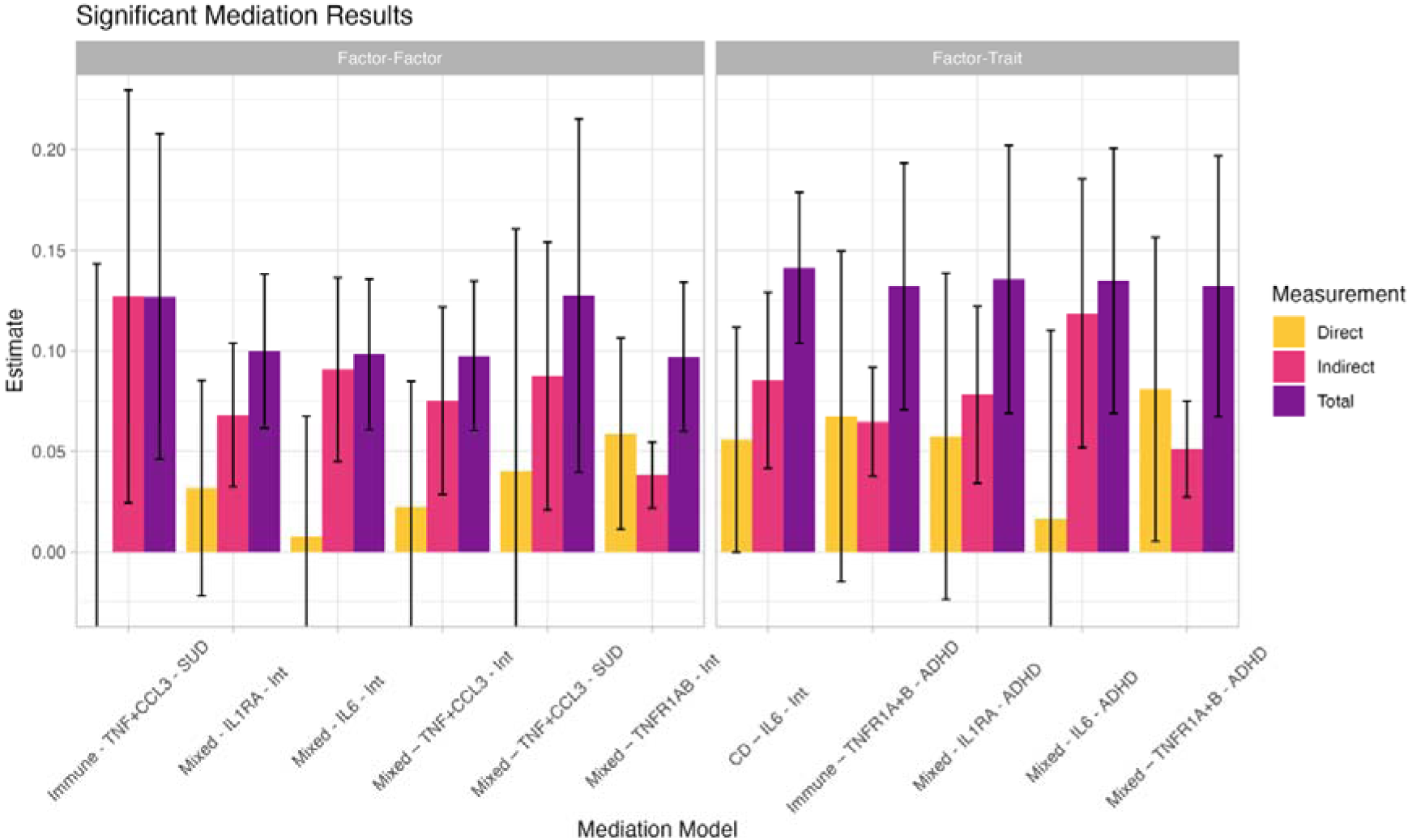
Significant Mediation Model Effect Estimates. Direct, indirect, and total effect estimates for each of the tested mediation models. Error bars display 1.96 * standard error. All mediation models displayed here were significant, as indicated by the significant indirect effect estimates and additionally, all total effect estimates were significant. Immune, autoimmune diseases; Mixed, mixed immune pattern diseases; Int, internalizing disorders; SUD, substance use disorders; CD, Crohn’s disease; ADHD, attention-deficit/hyperactivity disorder; TNF+CCL3, TNF- α/CCL3; IL6, IL-6; IL1RA, IL-1RA; TNFR1A+B, TNFR1A/B.

**Figure 4.**
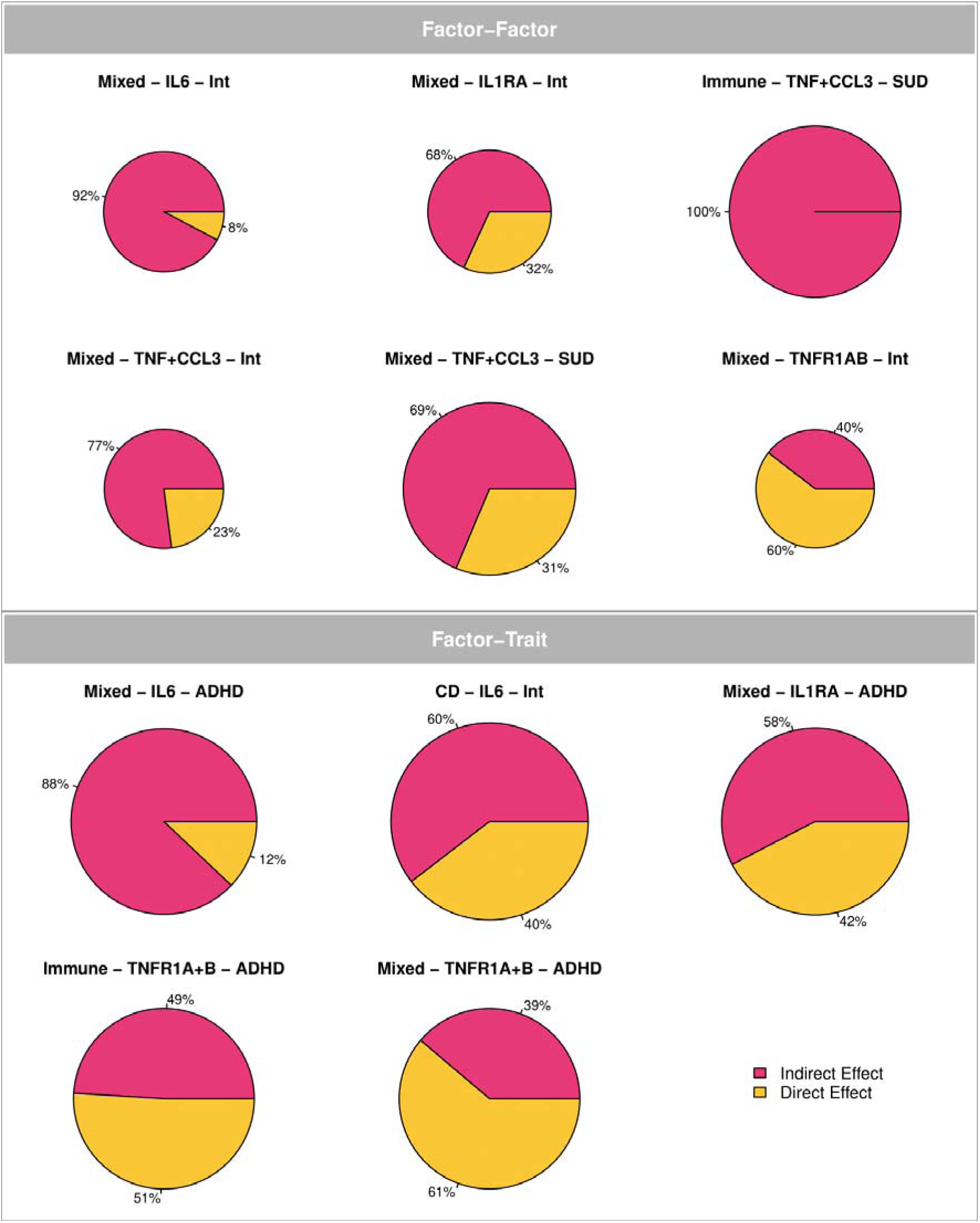
Proportions of Direct and Indirect Effects. Pie charts are displaying the proportions of the direct and indirect effects for each of the mediation models tested. The size of the pie charts represents the size of the total effect. Immune, autoimmune diseases; Mixed, mixed immune pattern diseases; Int, internalizing disorders; SUD, substance use disorders; CD, Crohn’s disease; ADHD, attention-deficit/hyperactivity disorder; TNF+CCL3, TNF-α/CCL3; IL6, IL- 6; IL1RA, IL-1RA; TNFR1A+B, TNFR1A/B. **A** displays all factor-factor relationships and **B** displays all factor-trait relationships.

Four of the six *factor-factor* models reflected mediation of the association between the mixed immune pattern and the internalizing factors by IL-6, IL-1RA, the combined TNF-α/CCL3 factor, and the TNFR1A/TNFR1B factor. The largest mediating effect was by IL-6, where the direct effect was attenuated to only .01 (*SE* = .03), and the indirect effect was .09 (*SE* = .02, *p_FDR_* = 2.98e-04), such that IL-6 mediated ∼92% of the total effect. The largest effect across models was for TNF-α/CCL3, which mediated 100% of the association between a substance use and autoimmune factor, wherein the direct effect was estimated at 0 (*SE* = .07), and the indirect effect at .13 (*SE* = .05, *p_FDR_* = 1.96e-02).

Within the *factor-trait* mediation models, we found that IL-6 mediated ∼60% of the relationship between the internalizing factor and Crohn’s disease, with a direct effect of .05 (*SE* = .02) and indirect effect of .09 (*SE* = .02, *p_FDR_*= 2.98e-04). Similar to the large number of mediation between mixed immune pattern diseases and internalizing disorders, three factor-trait mediation models were for the mixed immune pattern factor and ADHD, with the markers reflecting IL-1RA, TNFR1A/TNFR1B, and IL-6. The marker that again accounted for the largest proportion of the total effect was IL-6, with a direct effect of only .02 (*SE* = .05) and an indirect effect of .12 (*SE* = .03, *p_FDR_* = 7.97e-04) that accounted for ∼88% of the total effect.

### Multiple Mediator Models

The relationships between mixed immune pattern diseases and both internalizing disorders and ADHD were significantly mediated by several immune markers, which together accounted for more than 100% of the total effect. This, along with the fact that these markers are themselves highly genetically correlated (**Supplemental Figure 1)**, indicates that the mediating markers are tapping into interrelated biological pathways. This prompted running multiple mediator models that included the set of identified markers as correlated mediators (**Figure 5**). For the mixed immune pattern disease and the internalizing factor, the set of markers (IL-6, IL-1RA, TNF-α/CCL3, TNFR1A/B) collectively mediated 100% of the relationship, with a direct effect estimated at 0 (*SE* = .03, *p_FDR_* = 9.56e-01) and a significant collective indirect effect of all markers combined of .10 (*SE* = .02, *p_FDR_* = 2.79e-05).

**Figure 5.**
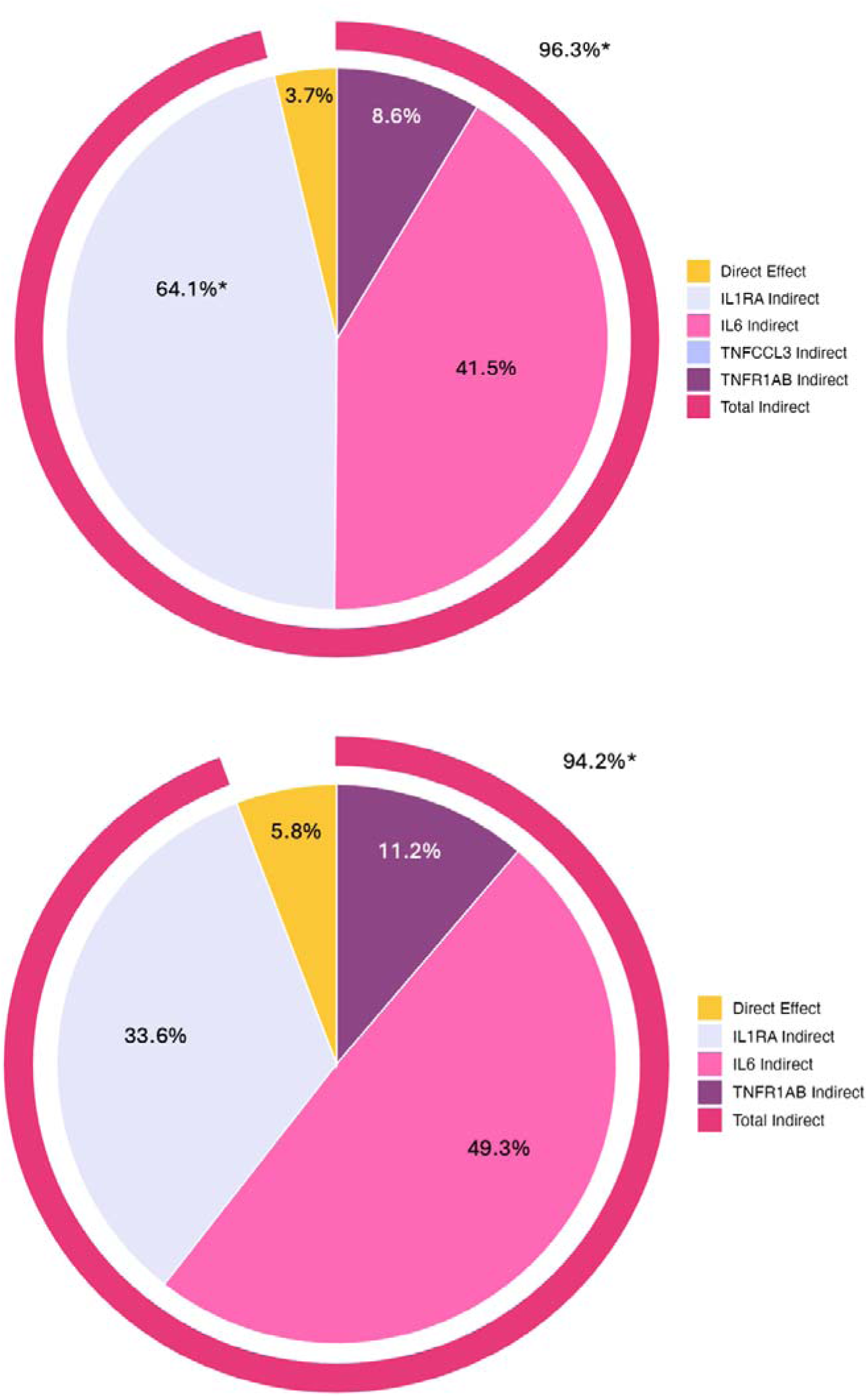
Multiple Mediation Results. Pie charts are displaying the proportions of the direct and indirect effects, with the total indirect effect broken down into the effects of the individual markers in a multiple mediation framework. Immune, autoimmune diseases; Mixed, mixed immune pattern diseases; Int, internalizing disorders; SUD, substance use disorders; CD, Crohn’s disease; ADHD, attention-deficit/hyperactivity disorder; TNF+CCL3, TNF-α/CCL3; IL6, IL-6; IL1RA, IL-1RA; TNFR1AB, TNFR1A/B. **A** Multiple mediation results for the mixed immune pattern disease and internalizing disorder relationship. **B** Multiple mediation results for the mixed immune pattern disease and ADHD relationship.

Only the individual mediating effect of IL-1RA remained significant with an indirect effect of .05 (*SE* = .02, *p_FDR_* = 1.75e-02). The multiple mediator model for ADHD by IL-1RA, IL-6, and TNFR1A/B revealed a non-significant direct effect (.01; *SE* = .05, *p_FDR_*= 8.84e-01) and a significant cumulative indirect effect of all markers combined of .13 (*SE* = .03, *p_FDR_* = 5.44e-06) that accounted for ∼94% of the total effect. We were not sufficiently powered to detect significant partial mediating effects of any markers.

## Discussion

Epidemiological and genetic work has described substantial relationships between immune-mediated diseases and psychiatric disorders. In the current study, we provide evidence that these genetic relationships are mediated by immune and inflammatory biomarkers. This represents a systematic and comprehensive multivariate solution to characterize the large number of bivariate associations that have been identified between immune-mediated and psychiatric diseases and immune markers. Our results clarify which of the genetic links between immune-mediated diseases and psychiatric disorders are mediated by immune markers, which markers mediate the relationships (and which markers do not), and which associations are due to biological processes unique to an individual trait relationships vs. more general to, for example, mixed immune pattern diseases and internalizing disorders as a class of diseases.

We found that the genetic relationships between immune-mediated diseases and psychiatric disorders are mediated by specific immune markers. Our analysis highlights the mediating roles of IL- 6, IL-1RA, and the shared biology across the TNF-α/CCL3 and TNFR1A/B factor. Notably, IL-6 significantly mediated the majority of three psychiatric-immune relationships: 92% of the association between internalizing and mixed immune pattern disease, 88% between ADHD and mixed immune pattern disease, and 60% of the genetic link between internalizing disorders and Crohn’s disease. The latter finding is consistent with prior conceptualizations of IL-6 as the inflammation-based mechanism underlying the association, which can lead from internalizing disorders to IBD^23^ and vice versa^24^. The broader set of associations for IL-6 suggest that it may confer more transdiagnostic risk beyond just the internalizing and Crohn’s relationship. At the same time, the TNF-α/CCL3 markers were found to significantly mediate 100% of the relationship between substance use disorders and autoimmune diseases and TNFR1A/B were the only markers that mediated the immune and ADHD association. In addition, the multiple mediator models for the mixed immune pattern and internalizing factors and ADHD revealed that the cumulative mediating effect across all markers was greater than the individual effect of all markers. These results collectively indicate the importance of multiple markers for psychiatric-immune relationships, though our results also help narrow the search space by identifying unrelated markers.

In addition to the significant associations observed, it is relevant to consider the patterns of null results, particularly given a literature that describes such widespread associations. For example, while our study links psychiatric disorders to IL-1β and TGFβ-1, they did not mediate any of the psychiatric-immune relationships. Furthermore, the included markers did not mediate the association between inflammatory diseases and the schizophrenia/bipolar factor, where the schizophrenia/bipolar factor was not associated with any of the markers, even though markers that were included here (like CRP and IL-6) have been previously associated with both disorders^25,26^. The most notable set of null results was for CRP, a marker that is associated with psychiatric disorders in addition to immune-mediated diseases^27–29^ and has shown significant overlap of genetic loci with those of psychiatric disorders^30^. This result may be surprising given the overwhelming number of studies linking CRP to psychiatric disorders^27^, with some going so far as to suggest using CRP as a biomarker for diagnosis psychiatric disorders and indicator of their severity^31^. The current findings revealed that the effect of CRP on immune-mediated diseases and psychiatric disorders was nonsignificant when IL-6 is a genetically correlated predictor. This is consistent with biological evidence, as CRP cannot cross the blood-brain-barrier (despite altering its permeability)^21^ while IL-6 can^22^, therefore making it more likely that IL-6 could have more direct links to psychiatric disorders whose genetic risk is concentrated in the brain. However, further follow-up on the mechanisms affecting this association is needed.

### Limitations and Future Directions

GWAS data was limited to individuals of European-like ancestry since it was only of sufficient sample size in this particular ancestral group. Future work should extend these analyses to other ancestral groups once sample sizes are sufficient. Furthermore, GWAS summary statistics included in this study remove the major histone complex (MHC) because of its complex LD-structure. However, the human leukocyte antigen gene (HLA)^32^ in the MHC region codes for genes that are relevant for immune function, and we are therefore missing genetic links that are driven by this region of the genome. Additionally, GWAS for biomarkers may include genetic signal for behavioral traits (such as smoking) that influence biomarker levels. Despite displaying the mediation models with directional arrows and specifying the models in terms of regression relationships, causal relationships between immune mediated diseases, psychiatric disorders, and immune markers cannot be inferred from our models. To gain a more holistic understanding of the mechanisms involved in the co-occurrence and genetic relationships between the diseases and immune functions and to potentially identify actionable treatment targets, future research needs to utilize the advantages of genetic tools such as Mendelian randomization to disentangle causal pathways underlying these relationships.

## Conclusion

Immune diseases and markers are widely associated with psychiatric risk, but comprehensive studies that evaluate the relative importance of different markers for a broad set of disorders are lacking. The present study provides insight into the potential mediating effects of 14 immune markers on genetic associations between factors defined by subsets of 11 immune-mediated diseases or 13 psychiatric disorders. Findings revealed null associations for CRP when including IL-6 as a correlated predictor. Six markers mediated different psychiatric-immune associations, with particularly large and pervasive effects observed for IL-6, though which markers were most relevant was specific to the disorders included. This indicates that the links across the psychiatric-immune space may include multiple biological pathways with some diagnostic specificity. Taken together, these findings elaborate on the etiology of the well-established links between immune diseases and psychiatric disorders, warranting further research into the underlying causes and temporal relationships of these associations.

## Methods

### Phenotype Selection and Quality Control

Recent studies of markers of the immune system associated with psychiatric disorders were referenced to compile an initial list of immune markers that was then filtered based on availability of sufficiently powered (reflected by a SNP-based heritability (h^2^) *Z*-statistic < 4^33^), publicly available genome-wide association study (GWAS) summary statistics. For each phenotype, we chose the most well-powered, publicly available GWAS conducted on unrelated individuals of European-like genetic ancestry. We excluded traits with GWAS summary statistics that had low genomic coverage (fewer than 850,000 SNPs), which led to a final selection of 14 immune markers. For psychiatric and immune-mediated disease traits, the same GWAS summary statistics of 13 disorders^17^ and 11 immune mediated diseases^4^ from their previously established factor structures were used respectively. In the case of the psychiatric disorders, the factor structure from the original paper was retained, while GWAS summary statistics that had a more recent version with larger sample sizes since the publication, were updated in the model. These included PTSD that increased in the total sample size (including cases and controls) from 9,537 to 1,254,808, bipolar disorder that increased from 413,466 to 840,309, and MDD with a sample size increase from 500,199 to 2,009,702. All GWAS summary statistics were subjected to a standardized quality control procedure using the "munge" function from the GenomicSEM R package. This function limits the data to HapMap3 SNPs, standardizes the reference alleles across all GWAS summary statistics, and applies filters for a minor allele frequency greater than 1% and imputation quality above 0.9, where available.

### LD-score Regression

We performed multivariable Linkage Disequilibrium (LD)-score regression (LDSC)^33^ using LD scores calculated from the European-like genetic ancestry subset of the 1000 Genomes Phase 3 project. The MHC region was excluded due to its complex LD structure, which could bias the results. Multivariable LDSC generates two matrices: (*i*) a genetic covariance matrix with SNP-based heritabilities on the diagonal and genetic covariances on the off-diagonal, and (*ii*) a sampling covariance matrix with squared standard errors of the estimates and sampling dependencies representing participant sample overlap on the diagonal and off-diagonal, respectively. For binary traits, such as disease or disorder status, the LDSC estimates were converted to a more interpretable liability scale, which accounts for the continuous distribution of genetic risk (liability) and corrects for sample ascertainment. The conversion to the liability scale was based on population prevalence from the corresponding GWAS paper or a representative prevalence for the study sample. Additionally, we used the sum of effective sample sizes from the contributing cohorts for the ascertainment correction, to produce the accurate estimates of the heritability on the liability-scale^34^. For these analyses, we entered a sample prevalence of 0.5 in LDSC, as the sample ascertainment had already been corrected through the effective sample size calculation. For continuous traits, such as the immune markers, the total continuous sample sizes were used, and no corrections were necessary.

### Genomic SEM

We utilized genomic SEM to model the genetic overlap across immune markers and subsequently conduct mediation analyses with immune markers mediating the genetic relationships between immune-mediated diseases and psychiatric disorders. Genomic SEM takes as input the output from multivariable LDSC along with a user specified SEM^35^. Good model fit was established based on the comparative fit index (CFI ≥ .9) and the standardized root mean squared residual (SRMR ≤ .1).

To determine the genetic factor structure of immune markers, we ran exploratory factor analyses (EFA) utilizing genomic E-SEM^36^. We specified for markers to have a standardized factor loading of at least .4. The best fitting was a three-factor solution that fit the data well (CFI = .959, SRMR = .071); however, this model excluded four of the markers that did not load onto any of the factors, and among the remaining 10 markers, many cross-loaded on multiple factors, indicating that the identified factor structure of the markers is non-parsimonious. To facilitate interpretation, individual markers of the immune system mediators of the relationships between immune disease and psychiatric disorders.

Before running the mediation models, we first tested whether the individual associations within the mediation model, denoted the *a*, *b*, and *c* path, were significant. The *a*, *b*, and *c* paths represent the immune-mediated disease factor to marker relationship, the psychiatric disorder to marker relationship, and the immune-mediated disease to psychiatric disorder relationship respectively. To do this, the five psychiatric and four immune disease factors were specified to correlate with all immune markers in two separate models. Significant correlations between immune disease factors and immune markers as well as between immune markers and psychiatric disorders factors denote significant *a* and *b* paths of the mediation model respectively. The *a* and *b* paths were considered as being significant if they surpassed the FDR-corrected significance threshold of 14x5 and 14x4 trait-marker combinations and if those relationships did not at the same time become *Q_Factor_* significant. *Q_Factor_* is a measure of local model misfit, specific to a pair of factors, that captures inter-factor correlations which fail to accurately reflect the genetic relationships between the indicators (in our case psychiatric disorders or immune mediated diseases) that define these factors^4^. When the correlation is significant and *Q_Factor_* is non-significant, it suggests that the factor correlation appropriately captures the broad pathways of risk sharing across these two clusters of disease traits. When *Q_Factor_* became significant for either the *a*, or the *b* path, we subsequently re-tested the other paths for all individual indicators on the factor to see which of the indicators on the tested factors remained significantly associated with markers and psychiatric disorders or immune-mediated diseases. Only mediation models for indicators with significant *a*, *b*, and *c* paths were then carried forward from the *Q_Factor_* significant factors.

This three-step process thus resulted in mediation models that tested a marker’s mediation of (i) an immune-mediated disease factor to psychiatric factor relationship (from here on denoted as *factor-factor* relationship), (ii) an immune-mediated disease factor to an individual psychiatric disorder relationship or a psychiatric disorder factor to and individual immune-mediated disease relationship (from here on denoted as *factor-trait* relationship), (iii) an individual immune-mediated disease to an individual psychiatric disorder relationship (from here on denoted as *trait-trait* relationship). Mediation models were set up and tested in genomic SEM by specifying “defined” parameters within the *lavaan* syntax for the total and indirect effect of the mediation, which has the advantage of providing standard error on the effects. Total effects were calculated as Total = *a* X *b* + *c’* and the indirect effects were calculated as Indirect = *a* X *b*.

### Multiple Regression

CRP and IL-6 showed identical correlation patterns with psychiatric and immune disease factors, leading to the same mediation models. Since IL-6 is a precursor of CRP and can cross the blood–brain barrier, unlike CRP, the CRP signal is likely explained by IL-6. This was tested by modeling multiple regressions in Genomic SEM to test the partial effects of both of those markers. In the case of one marker in a pair consistently exhibiting significant partial effects while the other one did not, we only proceeded to use the significant marker in the mediation models. All multiple regression results were FDR corrected for the number of regressions run for the marker pair.

### Multiple Mediators

Multiple psychiatric – immune relationships were significantly mediated by more than one immune marker. This result, alongside the strong genetic intercorrelations of the markers, indicates that some markers likely operate via shared biological pathways. To better understand the total mediation effect of all contributing markers combined, as well as the conditional mediating effects of each marker when accounting for correlated genetic signal, we fit a multiple mediator model to each of these immune-psychiatric relationships in Genomic SEM. For the multiple mediation models, the collective indirect effects were calculated as Indirect_Collective_ = (*a_k_* X *b_k_) +* (*a_j_* X *b_j_) +* (*a_i_* X *b_i_)* with *k*, *j*, and *i* denoting three different markers. The total effects were calculated as Total_Collective_ = *Indirect_Collective_* + *c’*.

## Data Availability

Summary statistics for data from the Psychiatric Genomics Consortium (PGC) can be downloaded or requested here: https://www.med.unc.edu/pgc/download-results/; Summary statistics for the Anxiety phenotype can be downloaded here: https://drive.google.com/drive/folders/1fguHvz7l2G45sbMI9h_veQun4aXNTy1v; Summary statistics for Celiac Disease: http://ftp.ebi.ac.uk/pub/databases/gwas/summary_statistics/GCST90014001-GCST90015000/GCST90014442/; Summary statistics for Systemic Lupus Erythematosus: http://ftp.ebi.ac.uk/pub/databases/gwas/summary_statistics/GCST003001-GCST004000/GCST003156/; Summary statistics for Diabetes Type 1: http://ftp.ebi.ac.uk/pub/databases/gwas/summary_statistics/GCST90014001-GCST90015000/GCST90014023/; Summary statistics for Rheumatoid Arthritis: http://plaza.umin.ac.jp/~yokada/datasource/software.htm; Summary statistics for Crohns Disease: http://ftp.ebi.ac.uk/pub/databases/gwas/summary_statistics/GCST004001-GCST005000/GCST004132/; Summary statistics for Ulcerative colitis: http://ftp.ebi.ac.uk/pub/databases/gwas/summary_statistics/GCST004001-GCST005000/GCST004133/; Summary statistics for Juvenile idiopathic arthritis: http://ftp.ebi.ac.uk/pub/databases/gwas/summary_statistics/GCST90010001-GCST90011000/GCST90010715/harmonised/; Summary statistics for Addisons Disease: http://ftp.ebi.ac.uk/pub/databases/gwas/summary_statistics/GCST90011001-GCST90012000/GCST90011871/harmonised/; Summary statistics for Myasthenia gravis: http://ftp.ebi.ac.uk/pub/databases/gwas/summary_statistics/GCST90093001-GCST90094000/GCST90093061/; Summary statistics for Psoriasis: http://ftp.ebi.ac.uk/pub/databases/gwas/summary_statistics/GCST90019001-GCST90020000/GCST90019016/; Summary statistics for Psoriatic arthritis: http://ftp.ebi.ac.uk/pub/databases/gwas/summary_statistics/GCST90243001-GCST90244000/GCST90243956/; Summary statistics for CRP can be downloaded here:https://ftp.ebi.ac.uk/pub/databases/gwas/summary_statistics/GCST90029001-GCST90030000/GCST90029070/ Summary statistics for the remaining immune markers can be downloaded or requested here:https://www.decode.com/summarydata/ Links to the LD-scores, reference panel data, and the code used to produce the current results can all be found at: https://github.com/GenomicSEM/GenomicSEM/wiki

