## Supplementary Figures for "Immune Markers Mediate Genetic Relationships between Immune Diseases and Psychiatric Disorders"

**Supplemental Figures**

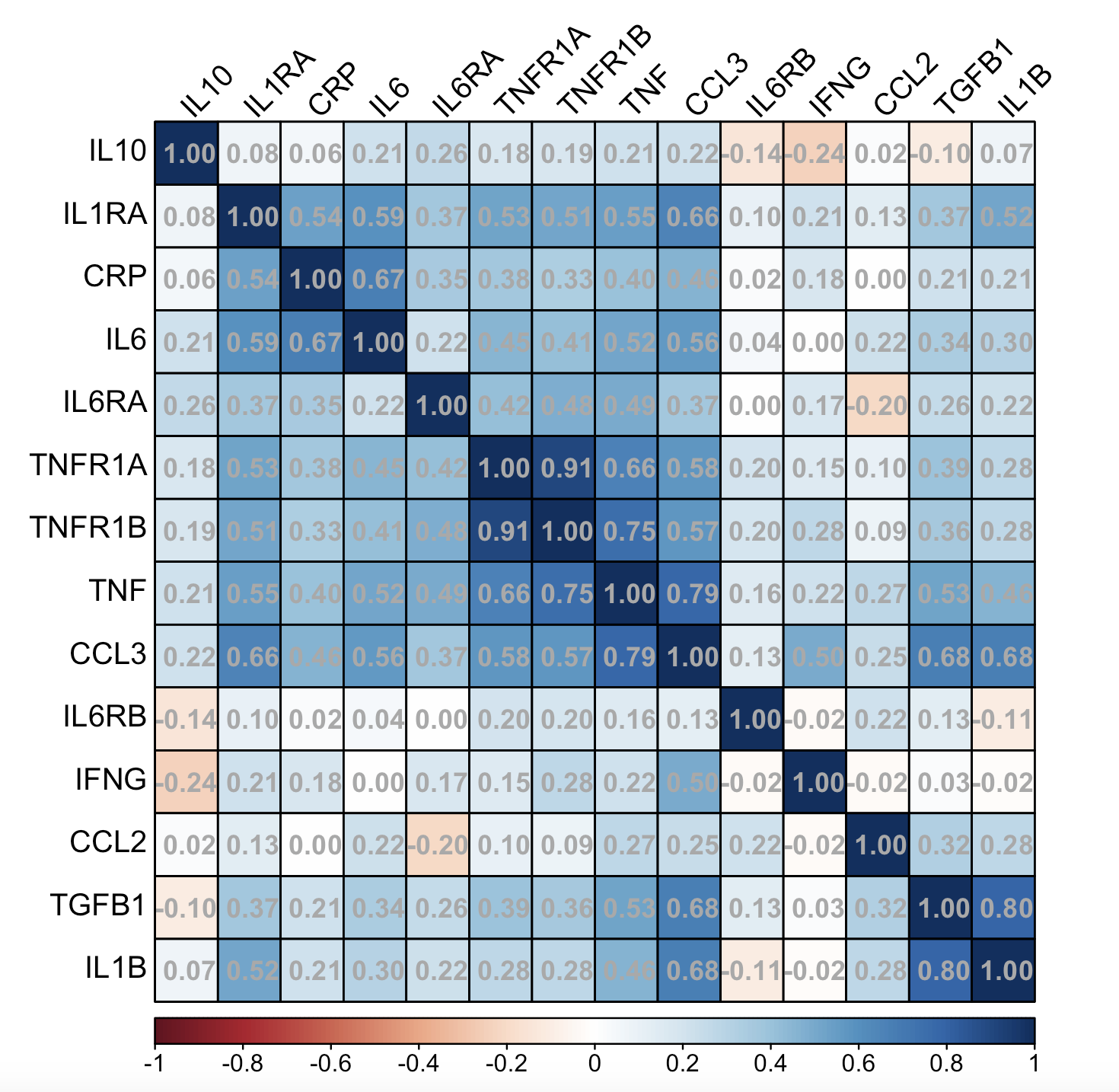

**Figure S1. Genetic Correlation Matrix.** Correlation matrix displaying genetic correlations between all included immune and inflammatory markers. Abbreviations for the markers can be found in Table 1.

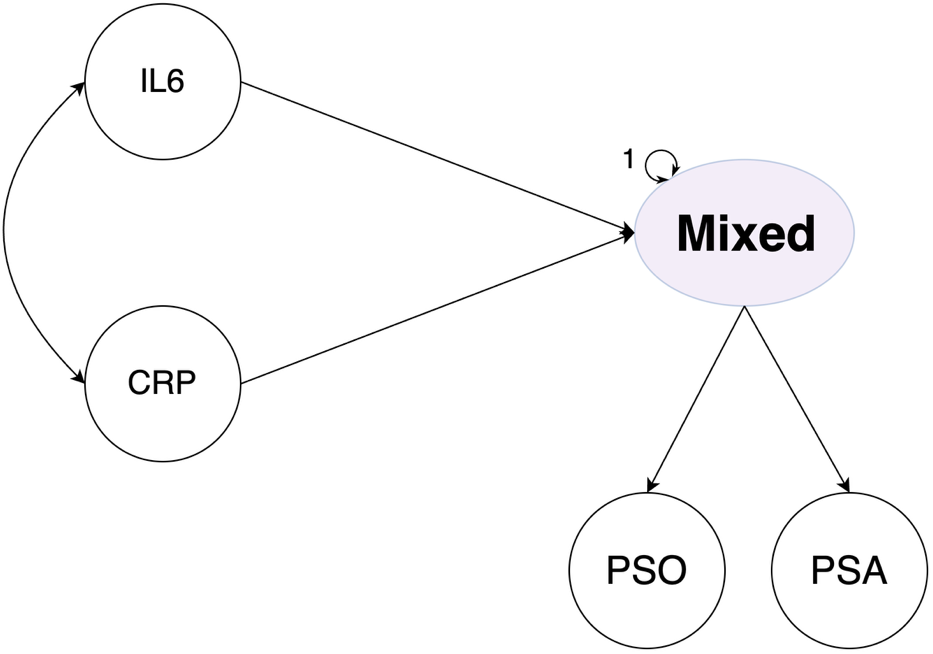

**Figure S2. Example Multiple Regression Model.** Example of a multiple regression model for the effects of CRP and IL6 on the mixed pattern immune disease factor.
